# Multi-omics analysis of renal clear cell carcinoma progression

**DOI:** 10.1101/2022.11.21.22282533

**Authors:** Anuj Guruacharya, James R Golden, Daniel Garrett, Deven Atnoor, Sujaya Srinivasan, Ujjwal Ratan, KT Pickard

## Abstract

Renal clear cell carcinoma (RCC), the most common type of kidney cancer, lacks a well-defined collection of biomarkers for tracking disease progression. Although complementary diagnostic and prognostic RCC biomarkers may be beneficial for guiding therapeutic selection and informing clinical outcomes, patients currently have a poor prognosis due to limited early detection. Without *a priori* biomarker knowledge or histopathology information, we used machine learning (ML) techniques to investigate how mRNA, microRNA, and protein expression levels change as a patient progresses to different stages of RCC. The novel combination of big data with ML enables researchers to generate hypothesis-free models in a fraction of the time used in traditional clinical trials. Ranked genes that are most predictive of survival and disease progression can be used for target discovery and downstream analysis in precision medicine. We extracted clinical information for normal and RCC patients along with their related expression profiles in RCC tissues from three publicly-available datasets: 1. The Cancer Genome Atlas (TCGA), 2. Genotype-Tissue Expression (GTEx) project, 3. Clinical Proteomic Tumor Analysis Consortium (CPTAC). Our study found that among others, gene expression levels (mRNA) from *GNG7* and *BCR* are potential predictors for RCC progression. For microRNA, we found hsa-mir-199a-2 and hsa-mir-129-1 to be potential predictors of RCC progression. Understanding how genes and protein expression levels change as RCC progresses will further guide the development of prognostic biomarkers and targets for RCC therapies.

## Introduction

Kidney cancer is among the ten most common cancers. In the US, rates for renal cell cancer continue to rise but mainly for early-stage tumors, with kidney renal clear cell carcinoma (RCC) accounting for 75% of all kidney cancers [1]. In the US, for the year 2020 the number of new cases of kidney cancer was 73,750 and deaths was 14,830 [2]. Although biomarkers for RCC are emerging [3, 4, 5, 6], patients usually have a poor prognosis due to limited early detection. Early identification of metastatic potential may be beneficial for therapeutic selection, as well as informing clinical outcomes. Using integrated diagnostic techniques made possible by lower-cost sequencing and predictive modeling, a more precise treatment approach may emerge by combining multimodal data such as mRNA, microRNA, and protein expression with clinical information [7, 8, 9, 10, 11].

Cancer databases contribute greatly to research that explains the molecular mechanisms underlying tumorigenesis. For example, the Cancer Genome Atlas (TCGA) database [12] consists of over 20,000 primary cancer and matched normal samples spanning 33 cancer types. Similarly, the Genotype-Tissue Expression (GTEx) database [13] contains tissue-specific gene expression for 54 non-diseased sites across nearly 1,000 individuals. The National Cancer Institute’s Clinical Proteomic Tumor Analysis Consortium (CPTAC) consists of over 2,000 proteomes and facilitates the discovery of cancer-specific protein biomarkers that augment DNA and RNA sequencing [14]. Bioinformatics mining—computational methods that are complementary to clinical trials—enables researchers to quickly evaluate concepts and generate hypothesis-free models. With publicly-available datasets becoming increasingly available, bioinformatics mining has led to a burst of research activity related to computational cancer biomarker and target discovery.

Understanding how genes and protein expression change as RCC progresses can further guide the development of prognostic biomarkers and treatment planning for RCC patients. In this study, we investigated how mRNA, microRNA, and protein expression levels were associated with different stages of RCC patients, as well as to their overall survival outcomes. Ranked genes that are most predictive of survival and disease progression can be used for target discovery for drugs and biomarker discovery for clinical decision support. In contemporary practice, VEGF and VEGF receptors are the most common therapy targets in the clinical treatment of RCC [15].

In this paper, we extracted clinical information of normal and RCC patients along with their related expression profiles in RCC tissues. All TCGA patients were clinically and pathologically diagnosed with RCC, and tumors were classified by TCGA into Stages I to IV based on the Fuhrman nuclear grading system [16]. Without *a priori* biomarker knowledge or histopathology information, we used ML techniques to investigate how clinical and omic data could be combined to identify the pathological stage of the tumor based on normal, early (Stage I+II), and late (Stage III+IV) classes.

## Methods

### Data collection and preparation

Clinical and genomic data were obtained from the Cancer Genome Atlas (TCGA) -Kidney Renal Carcinoma (KIRC) project, Genotype-Tissue Expression (GTEx), and the Clinical Proteomics Tumor Analysis Consortium (CPTAC). The processed mRNA data was used following the approach of Wang et al that combined data sources from TCGA and GTEx [17]. TCGA was the single source for microRNA data. Protein data and associated clinical data for those patients were obtained from CPTAC. Disease progression stages were defined by the American Joint Committee on Cancer (AJCC). Overall survival, overall survival months, and AJCC-defined pathologic tumor stage were extracted from clinical data and joined with genomic and protein data.

A combination of clinical and genomic data was provided as input to ML methods that included a decision tree model (XGBoost) and an ensemble (AutoGluon) model, which were trained to predict disease progression (Stages I through IV) for each of these datasets. We carried out a stratified 70/30 train-test data split before experiment training. Tables 1 and 2 show distributions of patients for each tumor stage based on the mRNA, microRNA, and protein datasets. A large class imbalance exists in each dataset, which is noted by the “Percentage of cohort” column in each table.

**Table 1:** For the different molecule types, the distribution of patients obtained from TCGA and GTEx for mRNA; TCGA for microRNA; and CPTAC for proteins.

| **Molecule type** | **Class groupings** | **Tumor stage** | **Number of patients** | **Percentage of cohort** | **Percentage of cohort** |
| --- | --- | --- | --- | --- | --- |
| mRNA | Class 0 | Normal | 104 | 18.71% | 18.71% |
|  | Class 1 | Stage I | 232 | 41.73% | 50.54% |
|  |  | Stage II | 49 | 8.81% |  |
|  | Class 2 | Stage III | 101 | 18.17% | 30.76% |
|  |  | Stage IV | 70 | 12.59% |  |
|  |  | Total | 556 | 100.00% | 100.00% |
| microRNA | Class 0 | Normal | 71 | 22.68% | 22.68% |
|  | Class 1 | Stage I | 121 | 38.66% | 47.60% |
|  |  | Stage II | 28 | 8.95% |  |
|  | Class 2 | Stage III | 48 | 15.34% | 29.71% |
|  |  | Stage IV | 45 | 14.38% |  |
|  |  | Total | 313 | 100.00% | 100.00% |
| proteins | Class 0 | Normal | 84 | 43.30% | 43.30% |
|  | Class 1 | Stage I | 52 | 26.80% | 33.51% |
|  |  | Stage II | 13 | 6.70% |  |
|  | Class 2 | Stage III | 33 | 17.01% | 23.20% |
|  |  | Stage IV | 12 | 6.19% |  |
|  |  | Total | 194 | 100.00% | 100.00% |

**Table 2:**
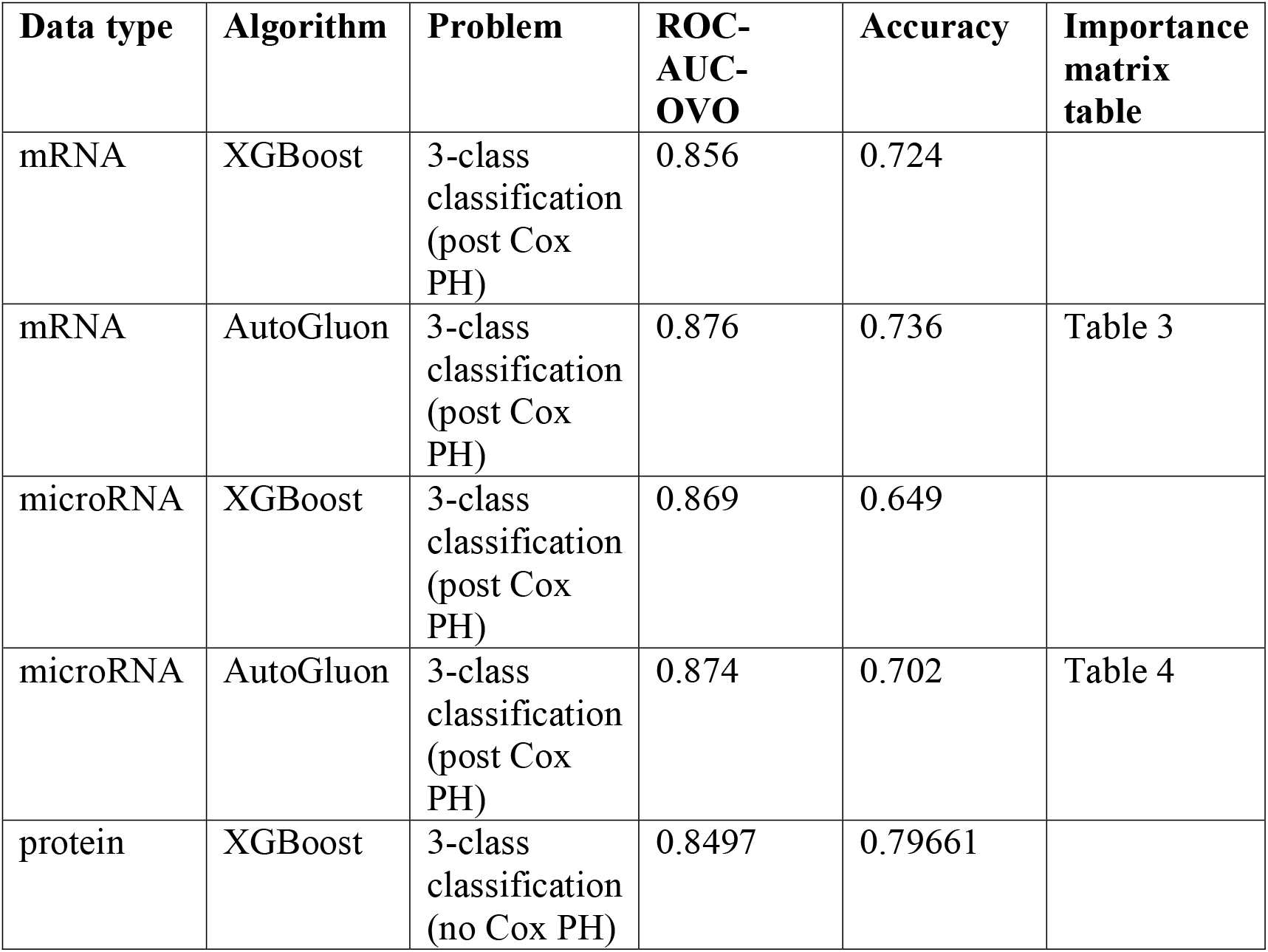
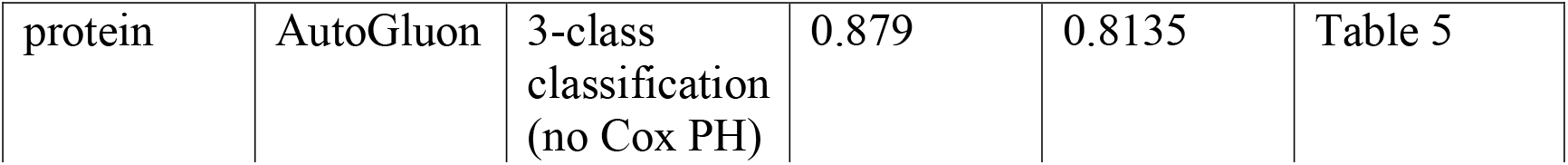
Comparison of evaluation metrics of the different models and methods used to predict stages.

| Data type | Algorithm | Problem | ROC-AUC-OVO | Accuracy | Importance matrix table |
| --- | --- | --- | --- | --- | --- |
| mRNA | XGBoost | 3-class classification (post Cox PH) | 0.856 | 0.724 |  |
| mRNA | AutoGluon | 3-class classification (post Cox PH) | 0.876 | 0.736 | Table 3 |
| microRNA | XGBoost | 3-class classification (post Cox PH) | 0.869 | 0.649 |  |
| microRNA | AutoGluon | 3-class classification (post Cox PH) | 0.874 | 0.702 | Table 4 |
| protein | XGBoost | 3-class classification (no Cox PH) | 0.8497 | 0.79661 |  |
| protein | AutoGluon | 3-class<br>classification<br>(no Cox PH) | 0.879 | 0.8135 | Table 5 |

**Table 3:**
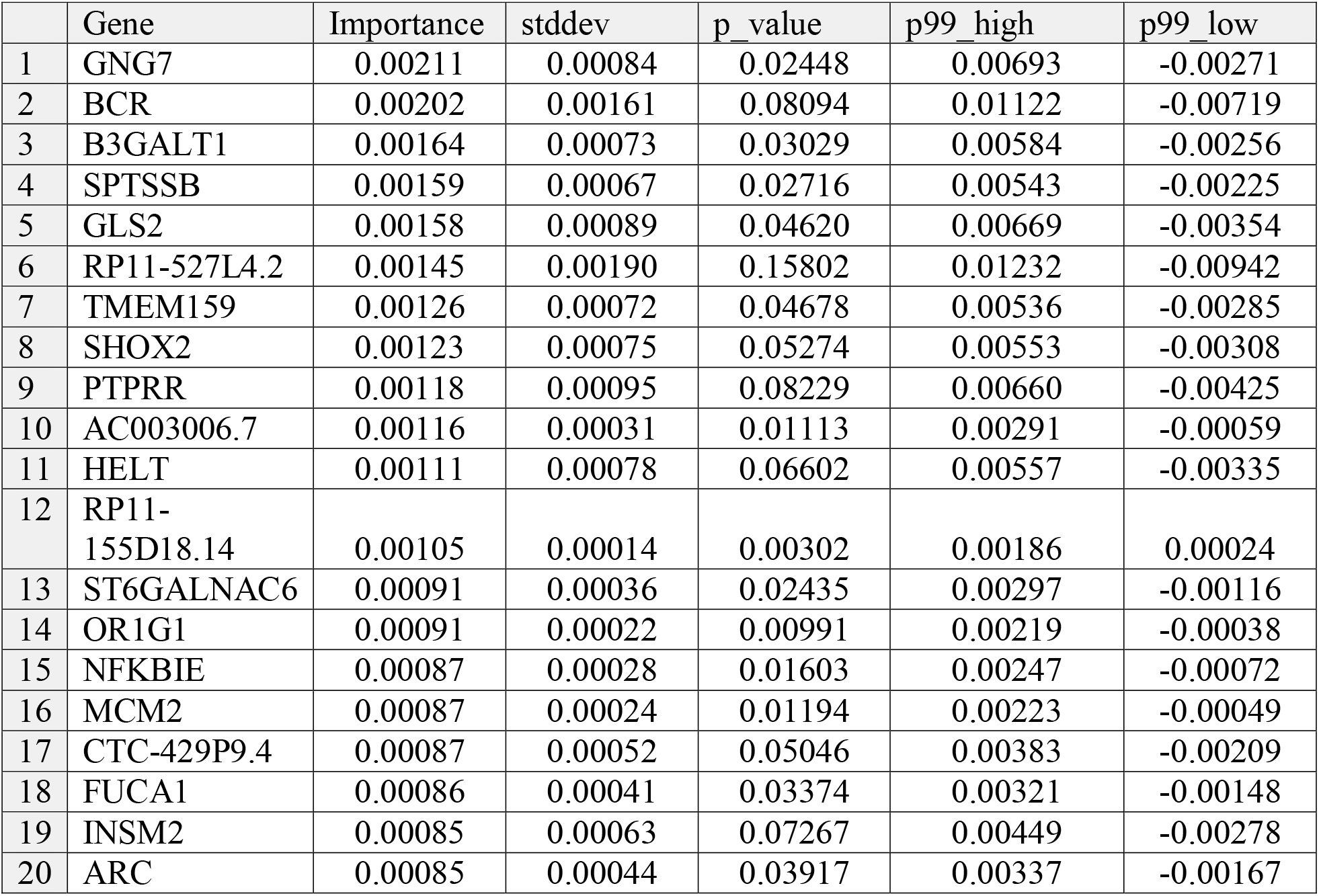
Top 20 genes ordered by importance in mRNA multi-class classification: AutoGluon.

|  | Gene | Importance | stddev | p_value | p99_high | p99_low |
| --- | --- | --- | --- | --- | --- | --- |
| 1 | GNG7 | 0.00211 | 0.00084 | 0.02448 | 0.00693 | -0.00271 |
| 2 | BCR | 0.00202 | 0.00161 | 0.08094 | 0.01122 | -0.00719 |
| 3 | B3GALT1 | 0.00164 | 0.00073 | 0.03029 | 0.00584 | -0.00256 |
| 4 | SPTSSB | 0.00159 | 0.00067 | 0.02716 | 0.00543 | -0.00225 |
| 5 | GLS2 | 0.00158 | 0.00089 | 0.04620 | 0.00669 | -0.00354 |
| 6 | RP11-527L4.2 | 0.00145 | 0.00190 | 0.15802 | 0.01232 | -0.00942 |
| 7 | TMEM159 | 0.00126 | 0.00072 | 0.04678 | 0.00536 | -0.00285 |
| 8 | SHOX2 | 0.00123 | 0.00075 | 0.05274 | 0.00553 | -0.00308 |
| 9 | PTPRR | 0.00118 | 0.00095 | 0.08229 | 0.00660 | -0.00425 |
| 10 | AC003006.7 | 0.00116 | 0.00031 | 0.01113 | 0.00291 | -0.00059 |
| 11 | HELT | 0.00111 | 0.00078 | 0.06602 | 0.00557 | -0.00335 |
| 12 | RP11-155D18.14 | 0.00105 | 0.00014 | 0.00302 | 0.00186 | 0.00024 |
| 13 | ST6GALNAC6 | 0.00091 | 0.00036 | 0.02435 | 0.00297 | -0.00116 |
| 14 | OR1G1 | 0.00091 | 0.00022 | 0.00991 | 0.00219 | -0.00038 |
| 15 | NFKBIE | 0.00087 | 0.00028 | 0.01603 | 0.00247 | -0.00072 |
| 16 | MCM2 | 0.00087 | 0.00024 | 0.01194 | 0.00223 | -0.00049 |
| 17 | CTC-429P9.4 | 0.00087 | 0.00052 | 0.05046 | 0.00383 | -0.00209 |
| 18 | FUCA1 | 0.00086 | 0.00041 | 0.03374 | 0.00321 | -0.00148 |
| 19 | INSM2 | 0.00085 | 0.00063 | 0.07267 | 0.00449 | -0.00278 |
| 20 | ARC | 0.00085 | 0.00044 | 0.03917 | 0.00337 | -0.00167 |

**Table 4:** Top 20 microRNA ordered by importance in multi-class classification selected: AutoGluon.

|  | gene | importance | stddev | p_value | p99_high | p99_low |
| --- | --- | --- | --- | --- | --- | --- |
| 1 | hsa-mir-199a-2 | 0.01996 | 0.00317 | 0.00415 | 0.03812 | 0.00179 |
| 2 | hsa-mir-129-1 | 0.01478 | 0.00533 | 0.02035 | 0.04532 | -0.01576 |
| 3 | hsa-mir-23b | 0.01195 | 0.00463 | 0.02333 | 0.03850 | -0.01461 |
| 4 | hsa-mir-200b | 0.01046 | 0.00566 | 0.04262 | 0.04290 | -0.02197 |
| 5 | hsa-let-7i | 0.00875 | 0.00867 | 0.11135 | 0.05845 | -0.04095 |
| 6 | hsa-mir-24-1 | 0.00525 | 0.00208 | 0.02441 | 0.01719 | -0.00670 |
| 7 | hsa-mir-142 | 0.00464 | 0.00353 | 0.07541 | 0.02488 | -0.01560 |
| 8 | hsa-mir-3676 | 0.00457 | 0.00494 | 0.12512 | 0.03288 | -0.02374 |
| 9 | hsa-mir-133b | 0.00450 | 0.00182 | 0.02521 | 0.01492 | -0.00592 |
| 10 | hsa-mir-3651 | 0.00413 | 0.00087 | 0.00727 | 0.00913 | -0.00087 |
| 11 | hsa-mir-188 | 0.00408 | 0.00372 | 0.09904 | 0.02541 | -0.01725 |
| 12 | hsa-mir-429 | 0.00400 | 0.00147 | 0.02109 | 0.01243 | -0.00443 |
| 13 | hsa-mir-139 | 0.00350 | 0.00231 | 0.06011 | 0.01676 | -0.00977 |
| 14 | hsa-mir-3200 | 0.00337 | 0.00425 | 0.15196 | 0.02775 | -0.02101 |
| 15 | hsa-mir-155 | 0.00320 | 0.00576 | 0.21852 | 0.03621 | -0.02981 |
| 16 | hsa-mir-3613 | 0.00305 | 0.00136 | 0.03031 | 0.01086 | -0.00476 |
| 17 | hsa-mir-3615 | 0.00297 | 0.00075 | 0.01033 | 0.00728 | -0.00133 |
| 18 | hsa-mir-130b | 0.00281 | 0.00445 | 0.19409 | 0.02830 | -0.02268 |
| 19 | hsa-mir-1307 | 0.00263 | 0.00064 | 0.00960 | 0.00631 | -0.00104 |
| 20 | hsa-mir-27b | 0.00225 | 0.00046 | 0.00696 | 0.00491 | -0.00041 |

**Table 5:** Top 20 genes ordered by importance in protein multi-class classification (AutoGluon) without survival analysis (from full set of available proteins).

|  | gene | importance | stddev | p_value | p99_high | p99_low |
| --- | --- | --- | --- | --- | --- | --- |
| 1 | MFSD4A_proteomics | 0.00859 | 0.00088 | 0.00172 | 0.01359 | 0.00357 |
| 2 | TUBB2B_proteomics | 0.00606 | 0.00473 | 0.07836 | 0.03317 | -0.02105 |
| 3 | TRIM37_proteomics | 0.00505 | 0.00437 | 0.09175 | 0.03011 | -0.02001 |
| 4 | CHL1_proteomics | 0.00479 | 0.00417 | 0.09232 | 0.02871 | -0.01911 |
| 5 | HLA-DRB1_proteomics_3 | 0.00455 | 0.00401 | 0.09425 | 0.02752 | -0.01842 |
| 6 | CRTC3_proteomics | 0.00434 | 0.00048 | 0.00205 | 0.00711 | 0.00157 |
| 7 | HLA-A_proteomics | 0.00354 | 0.00487 | 0.16777 | 0.03144 | -0.02437 |
| 8 | MT1A_proteomics | 0.00278 | 0.00219 | 0.07940 | 0.01531 | -0.00975 |
| 9 | HAPLN1_proteomics | 0.00202 | 0.00287 | 0.17340 | 0.01846 | -0.01441 |
| 10 | UTP11_proteomics | 0.00202 | 0.00116 | 0.04709 | 0.00865 | -0.00461 |
| 11 | PIGO_proteomics | 0.00202 | 0.00116 | 0.04709 | 0.00865 | -0.00461 |
| 12 | RFFL_proteomics | 0.00177 | 0.00044 | 0.00990 | 0.00427 | -0.00074 |
| 13 | DEFA3_proteomics | 0.00177 | 0.00044 | 0.00990 | 0.00427 | -0.00074 |
| 14 | COL2A1_proteomics | 0.00177 | 0.00306 | 0.21133 | 0.01931 | -0.01578 |
| 15 | HLA-DRB3_proteomics | 0.00177 | 0.00116 | 0.05904 | 0.00839 | -0.00486 |
| 16 | FTL_proteomics | 0.00177 | 0.00306 | 0.21133 | 0.01931 | -0.01578 |
| 17 | GSTZ1_proteomics | 0.00177 | 0.00044 | 0.00990 | 0.00427 | -0.00074 |
| 18 | MT1X_proteomics | 0.00177 | 0.00088 | 0.03641 | 0.00678 | -0.00324 |
| 19 | TFPI2_proteomics | 0.00177 | 0.00088 | 0.03641 | 0.00678 | -0.00324 |
| 20 | HBE1_proteomics | 0.00177 | 0.00088 | 0.03641 | 0.00678 | -0.00324 |

To increase potential clinical insight, patients were stratified into three distinct groups per dataset. These classes are defined as follows:

- Class 0: Normal Samples (includes matched normal samples)
- Class 1: Stage I and Stage II
- Class 2: Stage III and Stage IV

The stages of the samples were determined at the time of diagnosis by the TCGA consortium and are included in the public TCGA dataset. For this research, tumor stages were grouped together in classes based on biological similarities. For example, Stage I and II kidney tumors are concentrated within the kidney itself and combined into Class 1. Stage III and IV kidney tumors have progressed and spread outside of the kidney and combined into Class 2 [18]. This grouping allows for the identification of relevant potential biomarkers for tumor progression and the determination of patients with the highest risk of tumor progression. The distributions of patients over progression stages are shown in Table 1 by molecule type.

### Preprocessing

Patients with missing ground truth entries or genomic features with a high fraction of missing data were removed from the dataset. Genetic expression levels were normalized to zero mean and unit standard deviation.

### Survival analysis

The correlation between patient survival and tumor stage is shown in Table 2, which is supported by studies on genetics and survival [18]. Cox’s Proportional Hazard Test (Cox PH) was carried out for both mRNA and microRNA training sets separately based on the time to death in post-diagnosis days in order to determine which genes are significantly related to survival (p < 0.05) using the open-source Python package, *lifelines* [19]. Downstream models for disease progression were trained on a subset of each dataset containing only those genes significant for survival. This method allowed us to perform informed feature selection, reducing the mRNA feature space from 20,235 genes to 5,569 significant genes, and microRNA from 532 genes to 47 significant genes.

### XGBoost

The open-source Python package for XGBoost version 1.4.2 [20] was coupled with an open-source Bayesian-optimization package for hyperparameter tuning [22] over 50 rounds per model. A model with optimal hyperparameters was determined with 10-fold cross-validation on the training set and returned from a Bayesian optimization procedure with 1,000 boosting rounds. The final model with optimal parameter settings was trained on the full training set. Model performance was evaluated with accuracy and ROC-AUC-OVO (Area Under the Curve of the Receiver Operating Curve, using a One-Versus-One evaluation for imbalanced multi-class data) on the test set. We then used the XGBoost feature importance tool to create importance matrices.

### AutoGluon

AutoGluon version 0.3.1 [21], an open-source AutoML package, was also used to train a large ensemble model (linear, k-nearest neighbor, random forest, XGBoost, and tabular neural network models, among others) to compare with the custom XGBoost model, and also to compare the internal AutoGluon models that comprise the ensemble. The AutoGluon ensemble was trained for 30 minutes on each dataset, and, like the XGBoost models, the AutoGluon ensemble predictor was trained only on a subset of features deduced from Cox’s Proportional Hazard Test.

### Model Tuning

For hyperparameter tuning, we performed Bayesian optimization with 10-fold cross validation using the average ROC-AUC-OVO score as the evaluation metric. Optimization was accomplished using an open-source Python package, *BayesianOptimization* [22].

### Compute Environment

Models were trained using a ml.r5.4xlarge AWS SageMaker notebook instance. The code, models, and results are available in the GitHub repository listed in the Data Usage section. Data cleaning, Cox’s Proportional Hazard Test, XGBoost, and AutoGluon widely varied in time and computational resources due to the complexity of the data used (8,217 mRNA features as opposed to 11,710 protein features).

## Results

As described previously, mRNA and microRNA data were obtained from TCGA’s KIRC/RCC dataset, mRNA data for non-RCC patients from GTEx, and protein data (and cohort follow-up) from CPTAC. From the survival analysis, we found 8,218 mRNA, 38 microRNA, and 10 of the top 20 proteins that were significant (p < 0.05). These findings were used as classifier inputs.

Using mRNA, microRNA, and protein input data, we trained a classifier on a 70/30 stratified train-test split to predict three classes:

1. Class 0: Normal Patients
2. Class 1: Stage I and Stage II
3. Class 2: Stage III and Stage IV

On a test dataset of 30% of the data, we noted the evaluation metrics as in Table 2. AutoGluon outperformed XGBoost in every scenario, which is due to AutoGluon’s ensemble of models.

The mRNA, microRNA, and proteins found to be highly predictive of RCC progression are shown in Tables 2 through 5.

## Discussion and conclusion

### mRNA prognostic biomarkers

For mRNA gene expression potentially predictive of RCC progression, we will discuss the top 10 genes in descending order of importance (*GNG7, BCR, B3GALT1, SPTSSB, GLS2, RP11-537L4, TMEM159, SHOX2, PTPRR, AC003006*.*7*). *GNG7*, a modulator in various transmembrane signaling systems, had the highest ranking. *GNG7* is part of the guanine nucleotide-binding protein (G protein) family with downregulation associated with pancreatic and esophageal cancer [23, 24], as well as squamous cell carcinoma of the head and neck [25]. A further study by Xu et al found that *GNG7* contributes to the progression of clear cell renal cell carcinoma, which is in agreement with our findings [26]. *BCR* is a protein with two opposing regulatory activities toward small GTP-binding proteins, and has been associated with leukemia [27]. Studies by Kim et al have shown a relationship between BCR gene expression and RCC [28, 29]. Gene expression of *BCR* was also upregulated in KIRP, a renal cancer closely related to RCC [30]. *B3GALT1* is involved in the biosynthesis of the carbohydrate moieties of glycolipids and glycoproteins, and has been found to be involved in cervical adenosquamous carcinoma [31]. *SPTSSB* (serine palmitoyltransferase small subunit B) downregulation has been found to be detrimental to RCC patients [32]. *GLS2* promotes mitochondrial respiration and increases ATP generation in cells. In a study by Shi et al, *GLS2* was found to be upregulated in ccRCC cells; cells with *GLS2* shRNA displayed lower survival, lower glutathione levels, and a high lipid peroxide level [33]. *RP11-527L4* is a long intergenic non-protein coding RNA more commonly known as *LINC01976*. The relationship of this ln-RNA has not been studied with respect to renal cancer and searches did not reveal studies regarding its function. *TMEM159* plays an important role in the formation of lipid droplets, but we were unable to find literature associating *TMEM159* expression with RCC progression. *SHOX2* is an important gene implicated in craniofacial, brain, heart, and limb development [34], and a methylation analysis of *SHOX2* has been performed for early-stage lung cancer [35]. Further, Jung et al found that *SHOX2* gene body methylation was positively correlated with mRNA expression in RCC tissues [36]. *PTPRR* was found to be related with multiple types of cancer, but searches did not find an association with RCC. This gene is a member of the protein tyrosine phosphatase (PTP) family. PTPs are known to be signaling molecules that regulate a variety of cellular processes including cell growth, differentiation, mitotic cycle, and oncogenic transformation. Lastly, *AC003006*.*7* is an uncharacterized protein, with one transcript having a KRAB-containing protein. We could not find literature regarding this protein, but include it for completeness.

### microRNA prognostic markers

After comparing the top 10 microRNA sequences with search results from the mirbase database [37], several sequence candidates were implicated in RCC. For example, hsa-mir-301a has been cited as a possible biomarker of metastatic RCC [38]. hsa-mir-129-1 has been found to be a biomarker for hepatocellular carcinoma [39, 40], but no known relationship with RCC has been reported in literature. Hsa-mir-23b has been associated with various cancer types and is a promising therapeutic target [41]. Its downregulation has also been found to be a predictor of hepatocellular carcinoma progression. For miR-27b, Ishihara et al found that it functions as a tumor suppressor of RCC [42]. Hsa-mir-200b downregulation has been found to suppress metastasis of RCC [43], and hsa-let-7i has been implicated in RCC due to its relationship with *TRIM* genes [44]. Hsa-mir-24-1 plays a role in several different types of cancer [45] and may be connected to lupus nephritis [46]. Although hsa-mir-142 and hsa-mir-3676 have not been implicated in renal cancer in the literature, our final candidate, hsa-mir-133b, has been associated with renal cancer through the JAK2/STAT3 pathway [47]. It has also been identified as a biomarker in other studies [48].

### Protein prognostic markers

Considering the top 10 proteins identified from AutoGluon’s importance matrix, we first searched for them in the Human Protein Atlas [50]. The Human Protein Atlas performed a detailed analysis of each gene available in open-source TCGA datasets across several different cancer types. From this search, we found that all of the top 10 were considered cancer-related genes. Of these genes, 4 were considered prognostic markers for renal cancer (*MFSD4A, CHL1, HAPLN1, UTP11*), although 6 genes were not previously described as prognostic (*TUBB2B, TRIM37, HLA-DRB1, CRTC3, HLA-A, MT1A*). Of the 6 non-prognostic genes for RCC, 3 were considered prognostic for other cancers (*TUBB2B, TRIM37, HLA-A*), 1 was found to be cancer enriched in liver cancer (*MT1A*), and the final 2 were not found to be associated with high cancer specificity (*HLA-DRB1, CRTC3*) [49].

MFSD4A is a protein within the major facilitator superfamily domain (MFSD) that focuses on glucose transmembrane transport, and UTP11 is a protein that processes pre-18S ribosomal RNA [51]. MFSD4A and UTP11, while noted as prognostic markers for renal cancer survival, did not have additional supporting literature beyond citations found in the Human Protein Atlas.

Although both of these genes were among the highest-ranked by the AutoGluon model’s importance matrix—the first and tenth, respectively—more research is required to explore the connection between these genes and RCC patient survival rates. CHL1 is an ATP-dependent DNA helicase that promotes genomic stability through DNA replication, DNA repair, and ribosomal RNA synthesis [51]. CHL1 was found to be a prognostic marker for renal cancer by multiple sources [50, 52], suggesting that under-expression can lead to lower survival rates in renal cancer patients. HAPLN1 is a protein that binds hyaluronic acid with proteoglycan monomers [51]. Other studies have found that high expression of HAPLN1 leads to lower survival rates in RCC patients [53].

### About CPTAC Survival Data for Clear Cell Renal Carcinoma Patients and Contro

The results from Cox’s Proportional Hazard test on the CPTAC dataset for feature selection found only 3 significant proteins out of 11,170 (a 99.97% reduction). Due to the small sample size of deceased patients (12/203) and low correlation between tumor stage and vital status in the CPTAC cohort, the performance of XGBoost and AutoGluon models was evaluated on the full feature set of proteins from CPTAC data. XGBoost and AutoGluon performance improved when using all available proteins compared to the small subset found from Cox’s Proportional Hazard Test.

## Conclusion

Although our findings may provide new perspectives in understanding the pathogenesis of RCC, some limitations exist in our study. The dataset size was relatively small, and results could be improved with more survival data for CPTAC patients. We used the AutoGluon permutation shuffling approach to assess feature importance. One limitation of permutation shuffling is that it can produce unrealistic inputs as a result of the shuffling procedure [54, 55]. This study is an exploratory analysis using computational techniques and further bench experimental studies are required for validation before clinical use.

In conclusion, the use of ML techniques enabled the evaluation of different models to predict RCC cancer progression and identify potential prognostic markers. Predicting cancer stages from RNA expression data may be beneficial for therapeutic selection, as well as informing clinical outcomes. This work will assist clinicians to inform patient prognosis, and can be extended with other multimodal data such as imaging and DNA methylation datasets. Employing graph-based neural network models could further elucidate the interaction of genes identified in this paper.

## Data Availability

All data produced in the present work are contained in the manuscript.

https://github.com/aws-samples/biomarker-discovery

## Data Usage

TCGA and GTEx data are from https://doi.org/10.6084/m9.figshare.5330575[17]

CPTAC data used in this publication were generated by the Clinical Proteomic Tumor Analysis Consortium (NCI/NIH) [14]. Data were accessed through the Python module cptac, PMID: 33560848 (https://github.com/PayneLab/cptac). Data from clear cell carcinoma (kidney) were originally published in PMID: 31675502.

The code for this study is in the GitHub repository: https://github.com/aws-samples/biomarker-discovery.

